# Triple-drug therapy with ivermectin, diethylcarbamazine and albendazole for the acceleration of lymphatic filariasis elimination in Kenya: Programmatic implementation and results of the first impact surveillance

**DOI:** 10.1101/2024.01.30.24301765

**Authors:** Sammy M. Njenga, Henry Kanyi, Collins Okoyo, Edward Githinji, Cassian Mwatele, Hadley S. Matendechero, Wyckliff P. Omondi, Patrick N. Gitahi, Chrispin Owaga, Joyce K. Onsongo, Katherine Gass

## Abstract

The World Health Organization (WHO) endorsed the use of triple-drug mass drug administration (MDA) regimen with ivermectin, diethylcarbamazine (DEC) and albendazole (commonly abbreviated as IDA) to accelerate the elimination of lymphatic filariasis (LF) as a public health problem in settings where onchocerciasis is not co-endemic. The National Programme for Elimination of LF (NPELF) in Kenya was among the first adopters of the IDA-MDA and two annual rounds were provided in 2018 and 2019 to the residents of Lamu County and Jomvu sub-County in the coast region. There is a need to determine efficient sampling strategies, indicators, and the appropriate population groups that could be used for the monitoring and evaluation of LF programs using IDA-MDA for the elimination of the disease as a public health problem. Two cross-sectional surveys were conducted at baseline in 2018 before IDA-MDA and an impact assessment 17 months after the second round of IDA-MDA. The reported epidemiological treatment coverage was at least 80% in all implementation units during each round of IDA-MDA. Blood samples were tested for circulating filarial antigens using commercial Filariasis Test Strips (FTS) and any individual that was found to be positive was tested again at night for the presence of microfilariae in finger prick blood smears using microscopy. The overall prevalence of LF infection was relatively low at the baseline survey with Jomvu having 1.39% (95% CI: 0.91, 2.11) and Lamu having 0.48% (95% CI: 0.21, 1.13). Significant reductions in LF prevalence were observed during the impact assessment after two annual rounds of treatment. The overall relative risk reduction (%) in LF prevalence following the two rounds of MDA with IDA was significant in both Jomvu (52.45%, Z = –2.46, P < 0.02) and Lamu (52.71%, Z = –1.97, P < 0.05). Heterogeneity, however, was observed in the LF prevalence reduction between random and purposive clusters, as well as between adult and child populations. The results of the endline impact survey offered strong evidence that it was safe to stop the IDA-MDA in the two EUs because transmission appears to have been interrupted. It is also important to implement a post-treatment surveillance system which would enable efficient detection of any recrudescence of LF infection transmission at a sub-evaluation unit level. Our findings show that IDA-MDA may be considered for acceleration of LF elimination in other settings where onchocerciasis is not co-endemic.

**Author summary:** Preventive chemotherapy with antifilarial drugs is currently the mainstay public health intervention recommended for the elimination of lymphatic filariasis in endemic countries. Two-drug regimen mass drug administration of albendazole plus either diethylcarbamazine or ivermectin has been used widely in most countries since the launch of the Global Programme to Eliminate Lymphatic Filariasis by the World Health Organization in 2000. Safety and efficacy clinical studies conducted in Papua New Guinea have shown that co-administration of three antifilarial drugs namely, ivermectin, diethylcarbamazine and albendazole is superior than the two-drug regimen diethylcarbamazine and albendazole in clearance of microfilariae from the bloodstream. Consequently, the triple-drug regimen was endorsed for accelerating efforts to eliminate lymphatic filariasis as a public health problem in areas where onchocerciasis is not co-endemic. The aim of this study was to assess the feasibility of delivering mass treatment with the triple-drug regimen under programmatic conditions. An operational research study was conducted to determine the optimal sampling strategies, indicator (or set of indicators), and study populations for monitoring and evaluation of lymphatic filariasis programs using the triple-drug regimen mass drug administration. Two rounds of mass treatment with the triple-drug regimen were given to the residents of Lamu County (comprising of Lamu East and Lamu West sub-counties) and Jomvu sub-County (in Mombasa) in November 2018 and November 2019. The programme epidemiological treatment coverage was at least 80% in each implementation unit during the two treatment rounds. The preventive treatment resulted in significant reductions in the prevalence of the disease with the number of positive cases being fewer than the threshold recommended for areas where transmission is likely to be no longer sustainable.

## Introduction

Lymphatic filariasis (LF), commonly known as elephantiasis, is a mosquito-borne neglected tropical disease (NTD) caused by infection with the filarial nematode species *Wuchereria bancrofti, Brugia malayi* or *B. timori* (1). The parasites are transmitted from person to person by mosquitoes belonging to the genera *Culex, Anopheles, Mansonia* and *Aedes*. Clinically, LF infection may be asymptomatic, but can also lead to acute and chronic conditions, including lymphoedema (swelling of the tissues, primarily the legs) and hydrocele (scrotal swelling). In 2000, the World Health Organization (WHO) launched the Global Programme to Eliminate LF (GPELF) as a public health problem (2, 3). The GPELF has two principal aims: (i) to interrupt LF transmission, and (ii) to manage morbidity and prevent disability. To interrupt transmission of LF infection, the GPELF recommends annual community-wide mass drug administration (MDA) of two-drug treatment regimen (albendazole plus either ivermectin or diethylcarbamazine) to entire at-risk populations for 4 to 6 years at adequate levels of coverage. Modelling studies have estimated adequate treatment coverage to be at least 65% of the total population in endemic areas (4, 5).

It is estimated that the GPELF resulted in substantial health and economic benefits during the first 15 years (6). Overall, an estimated 36 million clinical cases and on average 175 million disability-adjusted life years (DALYs) were potentially averted during the period. In economic terms, an estimated US$100.5 million was potentially saved through medical expenses, potential income loss, and costs to the health system.

Approximately 4 million people are at-risk for LF in Kenya, residing in six counties (23 sub-counties) of the coastal region, including Kilifi, Kwale, Lamu, Tana River, Mombasa, and Taita Taveta. Kenya started an MDA programme with diethylcarbamazine (DEC) plus albendazole in 2002 in the then Kilifi district and expanded to include Malindi and Kwale districts in 2003. The geographic coverage was progressively expanded over the years to include other districts in the coastal region except Mombasa city and its peri-urban areas. However, the programme faced financial and operational challenges which led to MDA being missed in some years and complete cessation after the 2011 MDA. Nonetheless, the non-consecutive rounds of MDA led to considerable reduction in the prevalence of LF (7). The MDA programme was restarted in 2015 and baseline population surveys confirmed that LF prevalence was still greater than the 2% antigenemia threshold in several areas (8), thus justifying the resumption of MDA campaigns.

Findings of clinical trials suggest that co-administration of the three drugs used to treat LF (ivermectin, DEC, and albendazole – IDA) results in improved efficacy for clearing *W. bancrofti* microfilariae (Mf), which is the transmissible stage of the filarial parasite, and sterilization of adult filarial worms (9–11). In 2017, following a thorough review process of data from safety and efficacy trials of the triple-drug regimen, the WHO recommended it as an alternative MDA regimen for the acceleration of LF elimination (12). The guidelines recommend the use of annual IDA in specific settings where onchocerciasis is not co-endemic with LF including places where MDA has not yet started, fewer than 4 effective MDA rounds have been administered, and where MDA results have been sub-optimal. Following the WHO’s formal approval and release of the alternative treatment guidelines, Merck and the MECTIZAN^®^ Donation Program (MDP) promised to work with the WHO, national LF programs and other stakeholders to develop a mechanism to enable for the deployment of any required additional donation (13). The announcement was a major enabler for the adoption of the IDA regimen by endemic countries. Computer simulation modelling suggests that the triple-drug regimen has the potential to dramatically accelerate the elimination of LF if epidemiological treatment coverage is high and systematic non-compliance is low (14).

Sentinel site surveys conducted throughout the coastal region in 2015 and 2016 identified Lamu East and Jomvu sub-counties as the sites with the highest prevalence of LF (6.3% and 6.7% antigenemia, respectively) (8). Lamu county had received one round of MDA in 2011 and additional two rounds in 2016 and 2017 using the two-drug regimen of DEC and albendazole, and all the three MDA rounds had achieved effective coverage (i.e., ≥65% of the total population was treated). Jomvu, however, had received two rounds of MDA (in 2016 and 2017), but none exceeded the 65% treatment coverage threshold. Due to the historically high prevalence of LF, the intermittent history of MDA and the sub-optimal coverage, both Lamu county and Jomvu sub-county were selected to receive two annual rounds of the triple-drug (IDA) preventive chemotherapy in order to accelerate LF elimination. For successful implementation of MDA with IDA, WHO recommends epidemiological treatment coverage of 80% and above, with 100% geographic coverage.

In evaluation units where two-drug MDA is given for 5 to 6 consecutive years and the LF infection reduced to the recommended thresholds, the WHO recommends post-MDA surveillance using transmission assessment surveys (TAS) among six– and seven-year-old children (15). Critical cut-off values of numbers of circulating filarial antigen (CFA) positive children are employed to determine whether transmission has been interrupted in the evaluation units. While the TAS has proven to be an effective tool for basing MDA stopping decisions under the standard two-drug regimens, it is unclear whether the target age group and epidemiologic target (<2% antigenemia in areas with *W. bancrofti* infections) are appropriate when IDA is used. Because IDA will result in an accelerated interruption of transmission, an operational research study was done with the goal of establishing the target populations, infection indicators, sampling strategies, and thresholds required to determine when it is safe to stop MDA with IDA. In this paper, we report on the NPELF experience delivering MDA with IDA in three implementation units in coastal Kenya. The study also presents an analysis of the baseline and endline survey results from the operational research study in two evaluation units following IDA implementation.

## Methods and study design

### Study sites

Two rounds of MDA with IDA were provided in November 2018 and November 2019 in three sub-counties on the Kenyan coast: two (Lamu East and Lamu West sub-counties) in Lamu County and the other one (Jomvu sub-County) in Mombasa County. For the IDA impact survey, Lamu County, which is comprised of a mainland and several islands and is situated on north coast of Kenya near the border with Somalia, was considered as a single Evaluation Unit (EU), due to its relatively low population. Jomvu sub-county formed the second EU and encompasses a mixture of urban, peri-urban and a few rural areas. In both EUs, the village served as the primary sampling unit (PSU) for the cluster sampling survey.

### Approach for MDA implementation

Due to the critical importance of achieving high treatment coverage for MDA with IDA, and the fact that the new triple-drug regimen required ingestion of more medicines than the two-drug regimen (DEC and albendazole) used previously, various strategies were implemented to bolster compliance and treatment coverage.

### Robust partnerships

Local and international collaborations played a pivotal role in supporting the program’s execution. Partners such as Evidence Action, The Task Force for Global Health’s NTD Support Center, and the END Fund provided essential logistical, technical, and financial support, thus contributing to the successful implementation of the program. Evidence Action with support from the END Fund provided fiscal management and technical assistance in the following areas: design and procurement of training materials including those for advocacy, communication, and social mobilization (ACSM); logistics for drug delivery; and budget management for county and sub-county training and sensitization. The Kenya Medical Research Institute (KEMRI) with support from the NTD Support Center at The Task Force for Global Health provided technical support for baseline surveys and follow-up monitoring and evaluation (M&E). The Africa Institute of Health and Development (AIHD) conducted formative research that influenced the review of ACSM materials and identified specific population segments with lower program engagement that may benefit from intensified mobilization. The description and results of the formative research will be reported elsewhere.

### Adverse event management and pharmacovigilance tools

Since clinical trials in Papua New Guinea had shown that MDA with IDA is superior in parasite clearance, it was anticipated that there would be a heightened risk of adverse events (AEs) which necessitated deployment of a robust pharmacovigilance framework. Collaboration with Kenya’s Pharmacy and Poisons Board (PPB) enabled the implementation of comprehensive pharmacovigilance tools, including reporting forms. Medications to help manage any AEs were procured and stocked, including cetirizine, paracetamol, and prednisolone. The AE forms were part of the training manuals and community drug distributors’ (CDD) handbooks. To ensure that any community members’ questions or concerns about the drugs could be promptly addressed, the PPB gave their hotline numbers to be used in case of the occurrence of serious adverse events (SAEs).

### Enhanced Community Engagement and Supervision

Community health extension workers (CHEWs) played a pivotal role in engaging communities through community meetings (commonly known as *baraza*) and supportive supervision of the CDDs. The number of individuals targeted by each CDD was reduced from 500 to 300, which allowed the CDD to spend more time in each household to answer any questions, ensure accurate dosing and recording, and to directly observe the medicines being swallowed. There was also closer supervision of CDDs, with one CHEW overseeing ten CDDs, compared to 20 in the conventional two-drug MDA. Additionally, the duration of the IDA-MDA campaigns lasted for six days, compared to five days in the conventional two-drug MDA. All of these adjustments allowed the implementers to pay closer attention to detail and assured community members of the safety and importance of taking the medicines. Frequent echo-messaging was done with the CHEWs and chiefs reminding them of the key LF and IDA-MDA information they needed to share with their constituents through intense social mobilization.

### Innovative Delivery Strategies to reach program targets

Special attention was given to areas that were historically challenging to access, addressing the needs of systematic non-compliers. To maximize treatment coverage, the program ventured beyond conventional strategies by directly administering MDA in industrial areas and utilizing boats to reach remote islands. Fixed posts, temporary fixed posts, and door-to-door distribution were among the treatment strategies employed. For the first time, MDA was taken directly to factories in the industrial areas. This was an innovative way to capture populations typically missed by MDA campaigns by meeting them where they usually spend their days. Boats were also hired to reach the far-off islands that were previously missed and branded as hard-to-reach areas.

### Effective CDD selection and health care workers training

The selection of CDDs was a crucial determinant of program success. By recruiting retired health professionals and individuals from private chemists, the program leveraged the community’s trust in these respected persons. Similar strategies were employed in gated communities to enhance acceptance among diverse populations. These individuals are well known to the community members as health professionals which encouraged uptake. In the gated communities, prior appointments had to be made and members of these communities were selected as CDDs to boost the trust of these ‘upper class’ individuals. Training of CHEWs and other sub-county implementation teams was centralized and conducted simultaneously for all the cadres in order to ensure that accurate information was provided, and to minimize loss of information about the new triple therapy down the cascade.

### Involvement of champions

The involvement of individuals directly affected by the disease as LF champions during the MDA campaigns added a human element to the program. The narratives of the LF champions were shared during program launches, encouraging community members to actively participate in the IDA-MDA. During the launch of the campaign in Lamu, for example, these individuals openly explained how their lives had been negatively affected by LF, and dispelled the commonly held belief that hydrocele surgery would particularly make the men infertile. The support of the top leadership in the implementation areas was also critical for ensuring the success of the campaigns. In Lamu, for example, the Governor graced the IDA-MDA campaign launch and openly swallowed the medicines (a total of 9 pills) which sent a strong message that the intervention is indeed safe.

### Survey design and sampling strategy

Cross-sectional epidemiological and entomological baseline surveys and follow-up IDA-MDA impact assessment were conducted in 30 and 5-10 random and purposive clusters (villages), respectively. This report, however, focuses on the population-based epidemiological survey arm for which the baseline survey was conducted in October-November, 2018 and the follow-up impact assessment in April – May, 2021. The village served as the primary sampling unit (PSU) in this study. A two-armed sampling strategy was employed to select random and purposive PSUs as described below.

### Random sampling

All PSUs in the EU were listed in geographical order along with their respective population sizes from which 30 PSU (aka ‘clusters’) were selected using probability proportionate to estimated size (PPES) sampling. An Excel tool was created to aid study investigators in the selection of the clusters.

### Purposive sampling

During the baseline survey, 5 clusters in each EU were selected to become the purposive PSUs, based on suspicion of high transmission of LF. The selection of the 5 purposive clusters was done in consultation with the NPELF officers and local health workers. During the impact assessment, however, up to 10 clusters were purposively selected. To maximize the chances of selecting sites with ongoing transmission during the impact assessment, the purposive clusters were chosen to be the 5 villages with the highest CFA prevalence in children 5-9 years of age in the baseline survey and up to 5 additional villages that had the greatest proportion of positive mosquito pools (if these sites were different from the initial 5 selected villages based on CFA prevalence in children). If there were fewer than 5 sites with CFA positive children, the CFA prevalence in adults was used to determine greatest risk of LF transmission for the purposive site selection.

### Human sample size

Since the mosquito vectors responsible for LF transmission in coastal Kenya belong to *Culex* and *Anopheles* species (16), the sample size was powered to enable the detection of a 2% CFA threshold for each of two age groups: (1) children aged 5-9 years old and other community members aged >10 years during the baseline survey, and (2) children aged 5-9 years-old and other community members aged ≥18 years during the endline impact assessment. The sample size of the 5-9 years-old children that was required for the random cluster survey in each area was selected to be similar to the sample sizes used for the WHO-recommended TAS survey (15). The target population and sample size were adjusted for the endline impact survey based on WHO’s interim guidance to use a 1% Mf threshold in adults (≥18 years) with 95% confidence to assess the impact of IDA (17). S1 and S2 Tables show the sample sizes that were required during each survey time point. In each survey, the individuals were enrolled to participate if they had lived in the areas for at least 12 months.

### Blood sampling and testing for LF infection

Written informed consent was obtained from all adults and parents or guardians of children below 18 years old. Ethical approval of the study protocols and informed consent documents was obtained from the Kenya Medical Research Institute Scientific and Ethics Review Unit (KEMRI/SERU Protocol No. 3721 and 4099). Blood sampling was done in two human populations: children (age 5-9 years) and older persons (age ≥10 and ≥18 years in baseline and endline surveys, respectively). The blood specimens were tested for CFA in the field using the Filariasis Test Strip (FTS) as recommended by the manufacturer (Alere, Scarborough, Maine, USA). Briefly, 75μl of finger prick blood was collected from each consenting participant into one of the capillary containers provided by the manufacturer and placed on the test pad. The test results were read and recorded at exactly 10 minutes. Additional blood specimen was collected from the same prick site for preparation of a dried-blood spot (DBS) specimen to be used for serological assays later. Any individuals found to be positive by the FTS test were followed up for a second blood specimen collection during hours of peak Mf circulation (9 pm – midnight). Approximately 60μl of the nighttime blood sample was used for thick blood smear microscopy and another 60μl for DBS preparation. The nighttime DBS was preserved for later detection of parasite DNA using a quantitative real-time polymerase chain reaction (qPCR) method.

### Data management and analysis

In the field, data were collected on smartphones and regularly uploaded to an electronic database using the open-source software Secure Data Kit (SDK). Data sets were stored on a SQL secure server. Field staff were trained on how to use the tools for data collection in the field. The investigators had access to view and download the data sets in real time using a secure password-protected website.

A primary goal of the data analysis was to determine whether the decrease in LF prevalence among children aged 5-9 years is consistent with TAS guidelines for stopping MDA, as compared to any decrease in FTS and Mf prevalence among adults. To address this primary study objective, the prevalence of CFA was assessed within each evaluation unit by each age group (5-9 years and ≥18 years) and the upper 1-sided 95% confidence limits calculated, assuming a hypergeometric distribution and taking into account survey design.

Infection prevalence was calculated and the 95% confidence intervals (CIs) determined using a binomial regression model, taking into account clustering by villages. To assess impact of IDA, the relative risk reductions (RRR) in CFA prevalence from baseline to endline impact survey were calculated using multivariable mixed effects models with random intercepts for the clusters and implementation units and P-values obtained using Wald test. The relative risk reduction was calculated using the following formula, 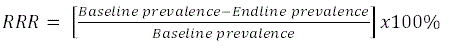.

All statistical analyses were carried out using STATA version 16.1 (STATA Corporation, College Station, TX, USA). Graphs were developed using the ggplot package implemented in R software. Village locations were mapped using ArcGIS Desktop version 10.2.2 software (Environmental Systems Research Institute Inc., Redlands, CA, USA).

## Results

In Kenya, the MDA implementation unit is the sub-county (equivalent of district in most other African countries). Although Lamu County was considered as a single EU for the operational research study, the two sub-counties in this area were considered separate implementation units (IUs) during MDA with IDA. Table 1 shows the epidemiological treatment coverage (reported coverage) with IDA-MDA in the three IUs in 2018 and 2019. An independent treatment coverage evaluation survey undertaken soon after the IDA-MDA to validate the epidemiological treatment confirmed that high treatment coverage was indeed achieved (Table 1).

**Table 1.**
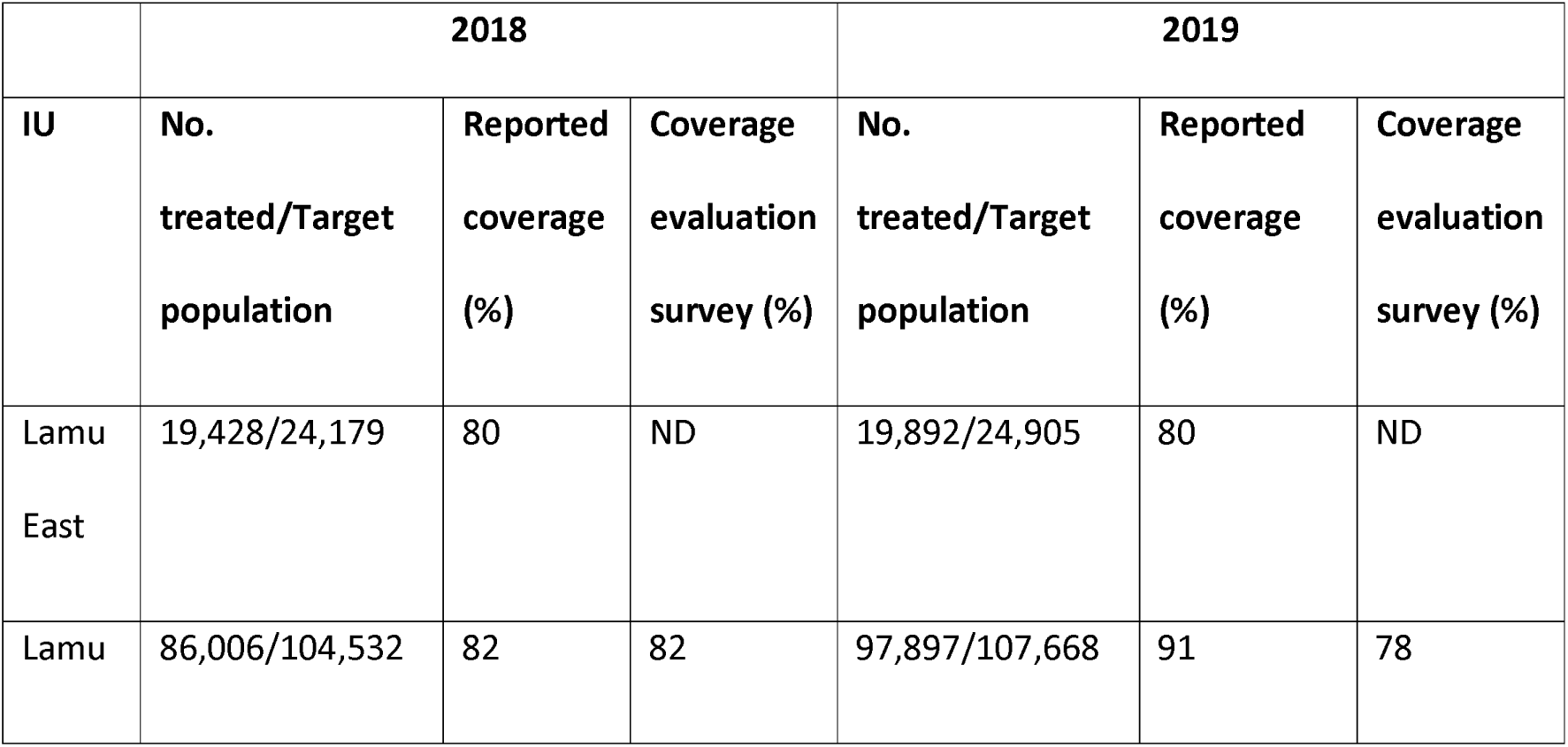
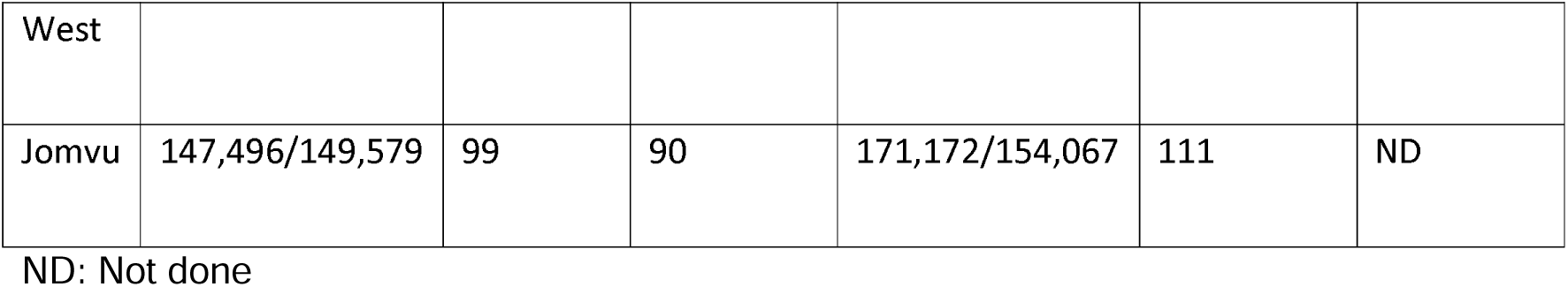
Coverage of two rounds of mass drug administration (MDA) with triple-drug therapy (ivermectin, diethylcarbamazine and albendazole – IDA) in Lamu and Jomvu implementation units, 2018 and 2019.

Overall, a total of 6,559 individuals were enrolled for the baseline survey out of whom 3,095 and 3,464 were from Lamu and Jomvu, respectively (Table 2). Out of 10,694 individuals enrolled during the IDA-MDA endline impact survey, 5,235 and 5,459 were from Lamu and Jomvu, respectively.

**Table 2.**
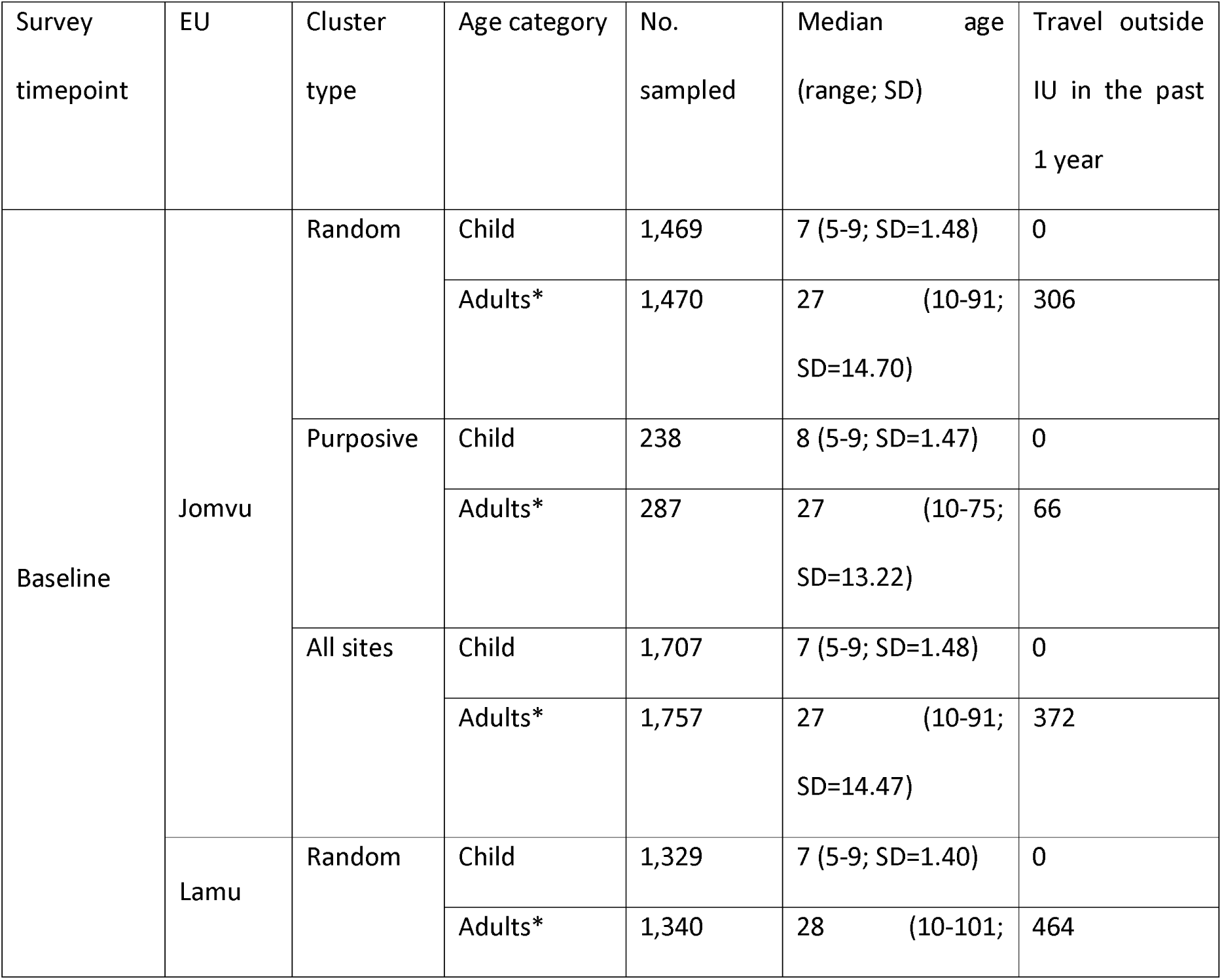
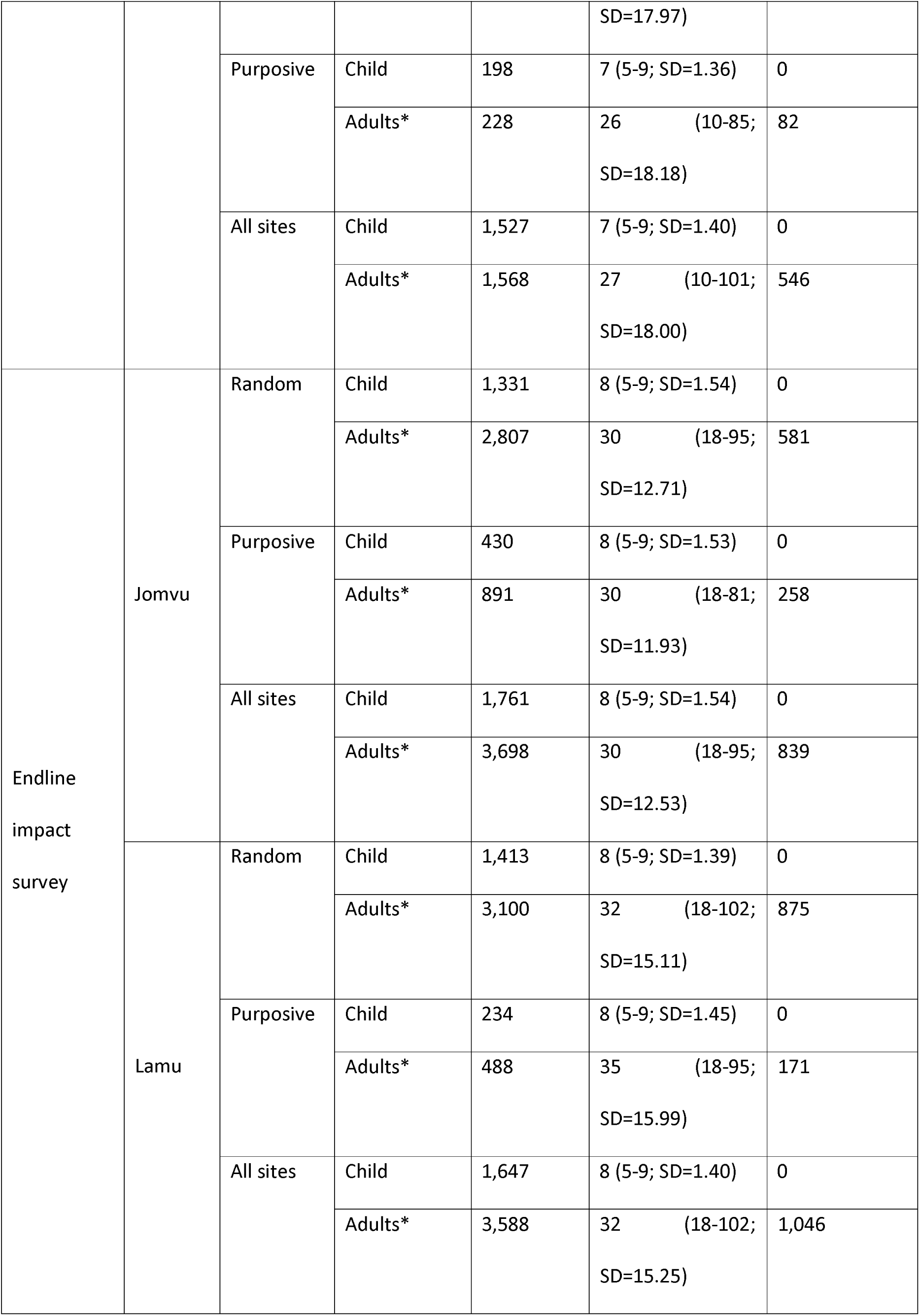
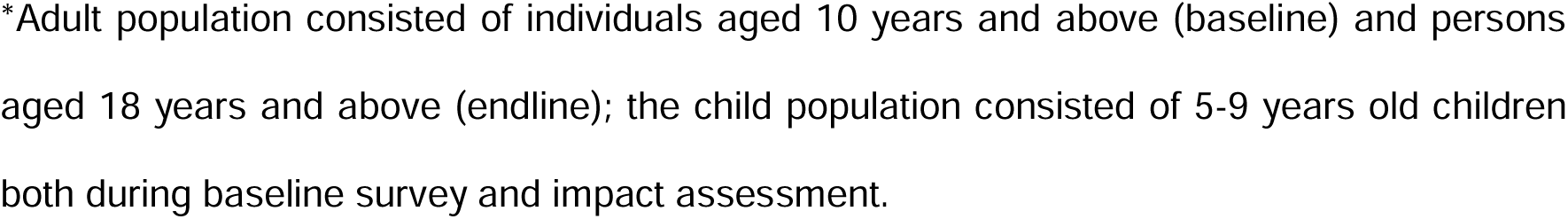
Baseline and IDA-MDA endline impact survey characteristics in each evaluation unit (EU).

Table 3 summarizes the prevalence of LF infection in the two EUs, while comparing children and adult populations and the random and purposive clusters. In general, the overall prevalence of LF infection by FTS in both types of clusters was relatively low at the baseline survey, with Jomvu having 1.39% (95%CI: 0.91,2.11) and Lamu having 0.48% (95%CI: 0.21, 1.13). Of 15 FTS positive individuals identified during the baseline survey in Lamu, 3 were children between the ages of 5 and 9 years. In contrast, of the 48 positive cases identified during the baseline survey in Jomvu, 11 were children aged 5-9 years. Of the 63 FTS positive cases, one individual aged between 40 and 50 years in Jomvu was found to have three (3) Mf in the night blood sample.

**Table 3.**
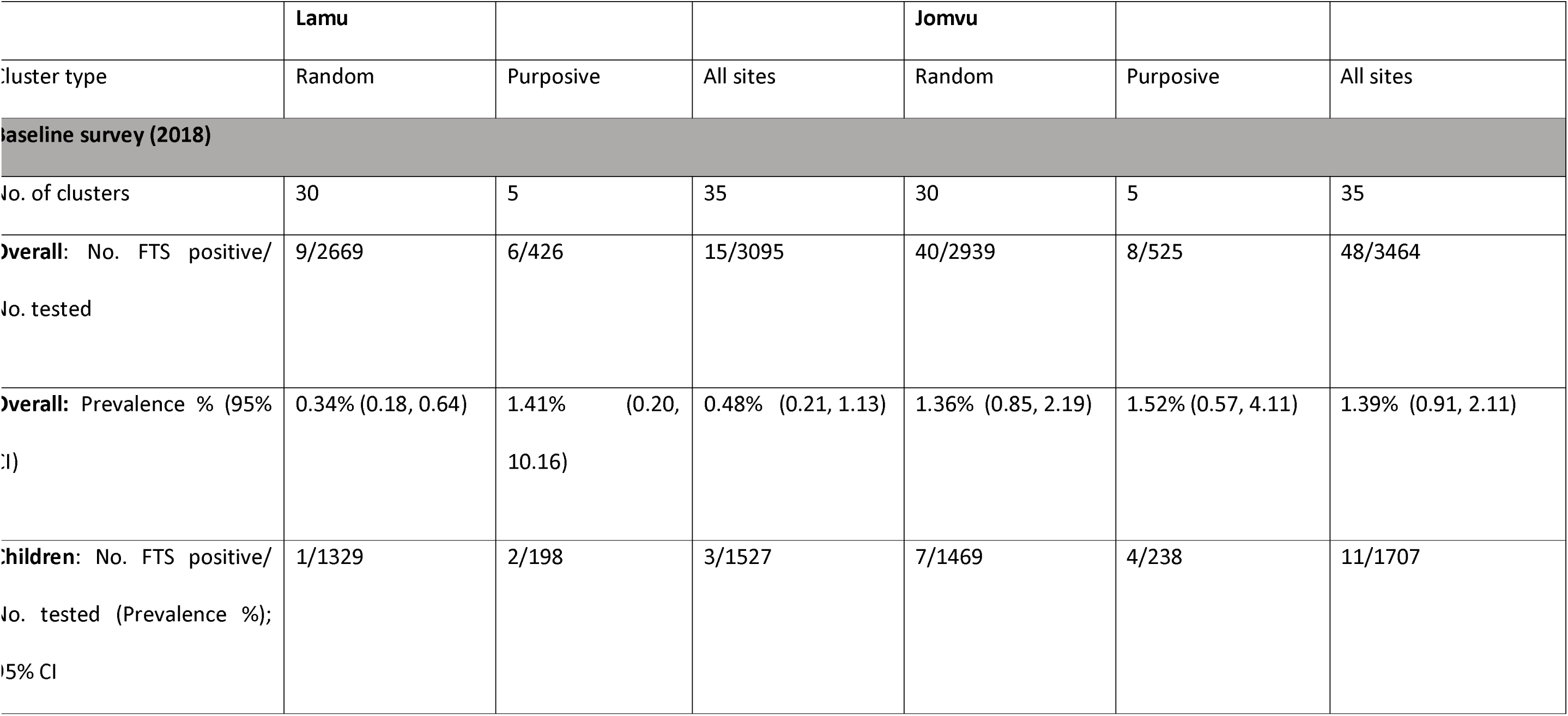
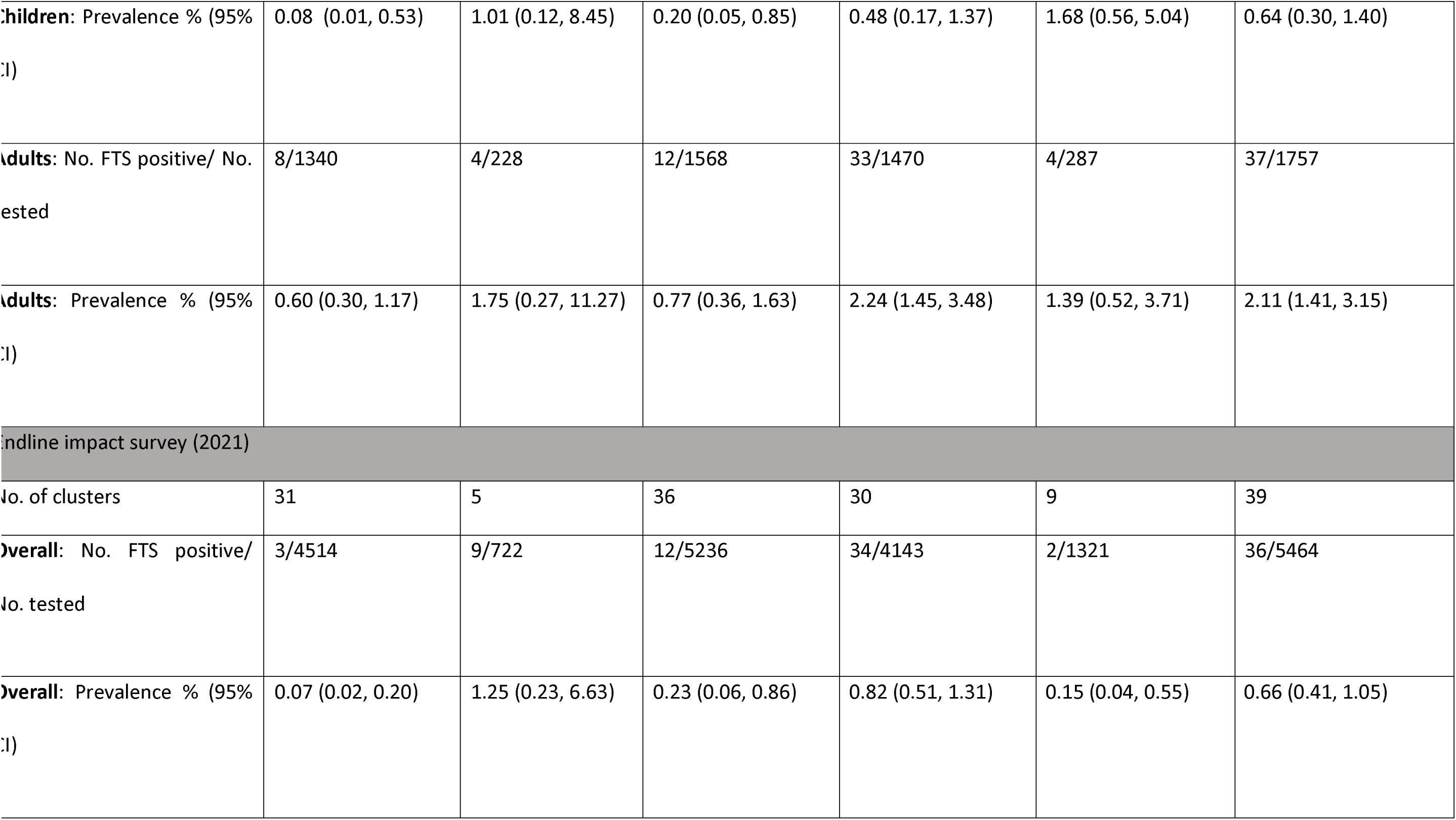
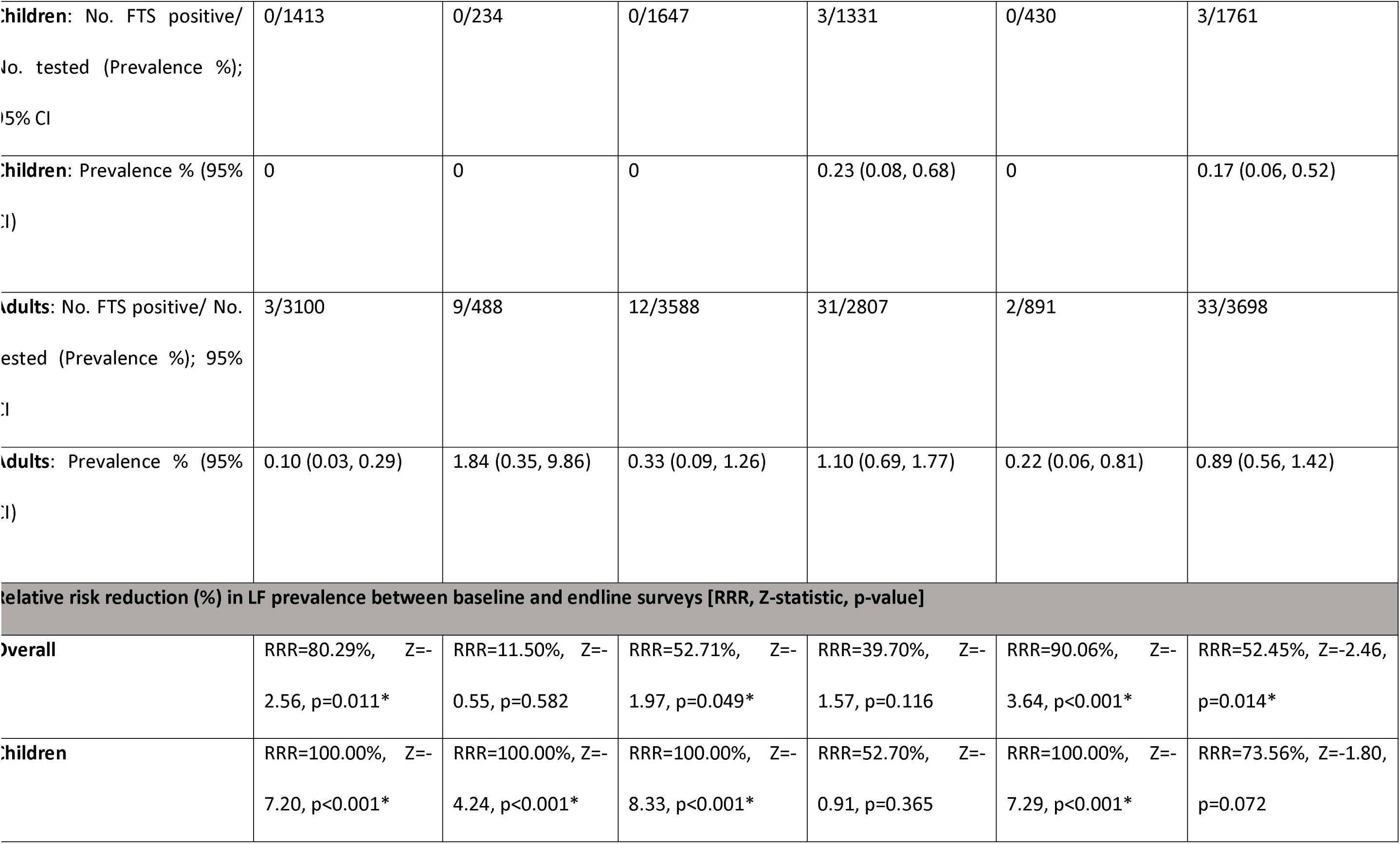
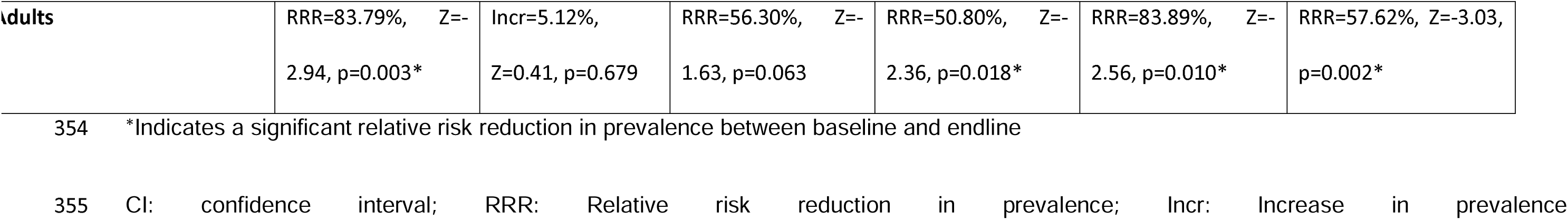
LF infection prevalence during baseline and endline surveys, and relative reduction (RR %) stratified by cluster type.

While there was an apparent reduction in the overall prevalence of LF infection following the administration of two rounds of MDA with IDA, the reduction was not consistent across the two EUs and study populations. Despite Lamu having very low numbers of LF positive cases in the baseline survey, there was observed statistically significant decrease in prevalence between the baseline and endline surveys, RRR = 52.71% (Wald test: Z = –1.97, p = 0.049). This significant decline in Lamu was largely attributed to the random clusters, RRR = 80.29% (Wald test: Z = –2.56, p = 0.011). In Jomvu, where LF prevalence was relatively higher at baseline survey, the overall reduction in LF prevalence was significant in the purposive villages, RRR = 90.06% (Wald test: Z = –3.64, p < 0.001), but not in the random sites, RRR = 39.70% (Wald test: Z = –1.97, p = 0.116). The reduction in LF prevalence among the adult population after the two rounds of IDA-MDA in Jomvu was significant in both random, RRR = 50.80% (Wald test: Z = –2.36, p = 0.018) and purposive, RRR = 83.89% (Wald test: Z = –2.56, p = 0.010) clusters. Restricting the analysis to children in Jomvu, the reduction was only significant among the purposive villages, RRR = 100.00% (Wald test: Z = –7.29, p < 0.001) but not in the randomly selected villages, RRR = 52.70% (Wald test: Z = –0.91, p = 0.365).

In general, there was a greater number of LF positive clusters in Jomvu compared with Lamu during both surveys (Table 4, and Figs 1 and 2). A total of 11 out of 35 clusters in Jomvu had LF prevalence of 2% or above during the baseline survey, compared with 4 out of 39 clusters during the endline impact survey. In contrast, Lamu had 2 out of 35 clusters with LF prevalence of 2% or above during the baseline survey and 1 out of 36 clusters during the endline impact survey.

**Fig 1.**
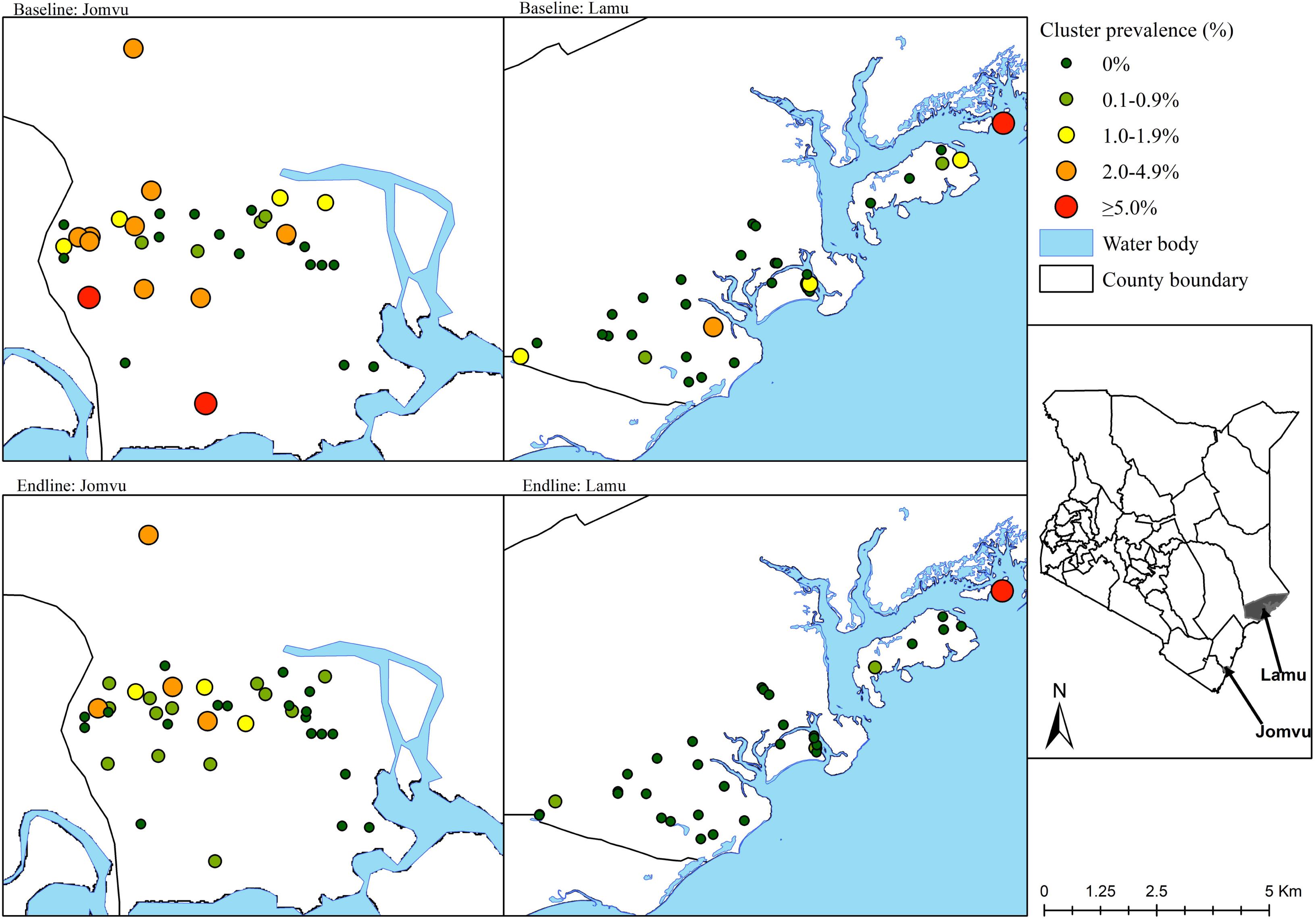
LF infection rates by FTS test at baseline and endline (after 2 rounds of IDA-MDA) among study clusters in the two EUs. The inset map of Kenya shows the location of the two study areas.

**Fig 2.**
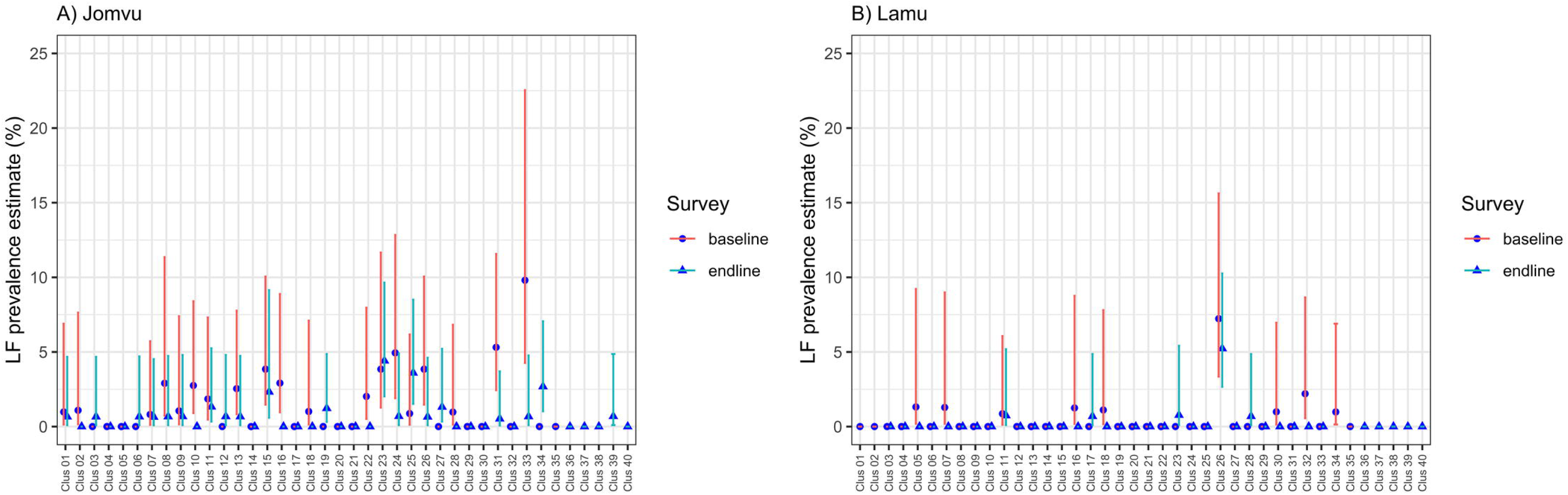
Village-level prevalence of LF infection by FTS test at baseline and endline (after 2 rounds of IDA-MDA), stratified by evaluation unit. Points represent point prevalence (%) with 95% confidence interval (CI) error bars.

**Table 4.**
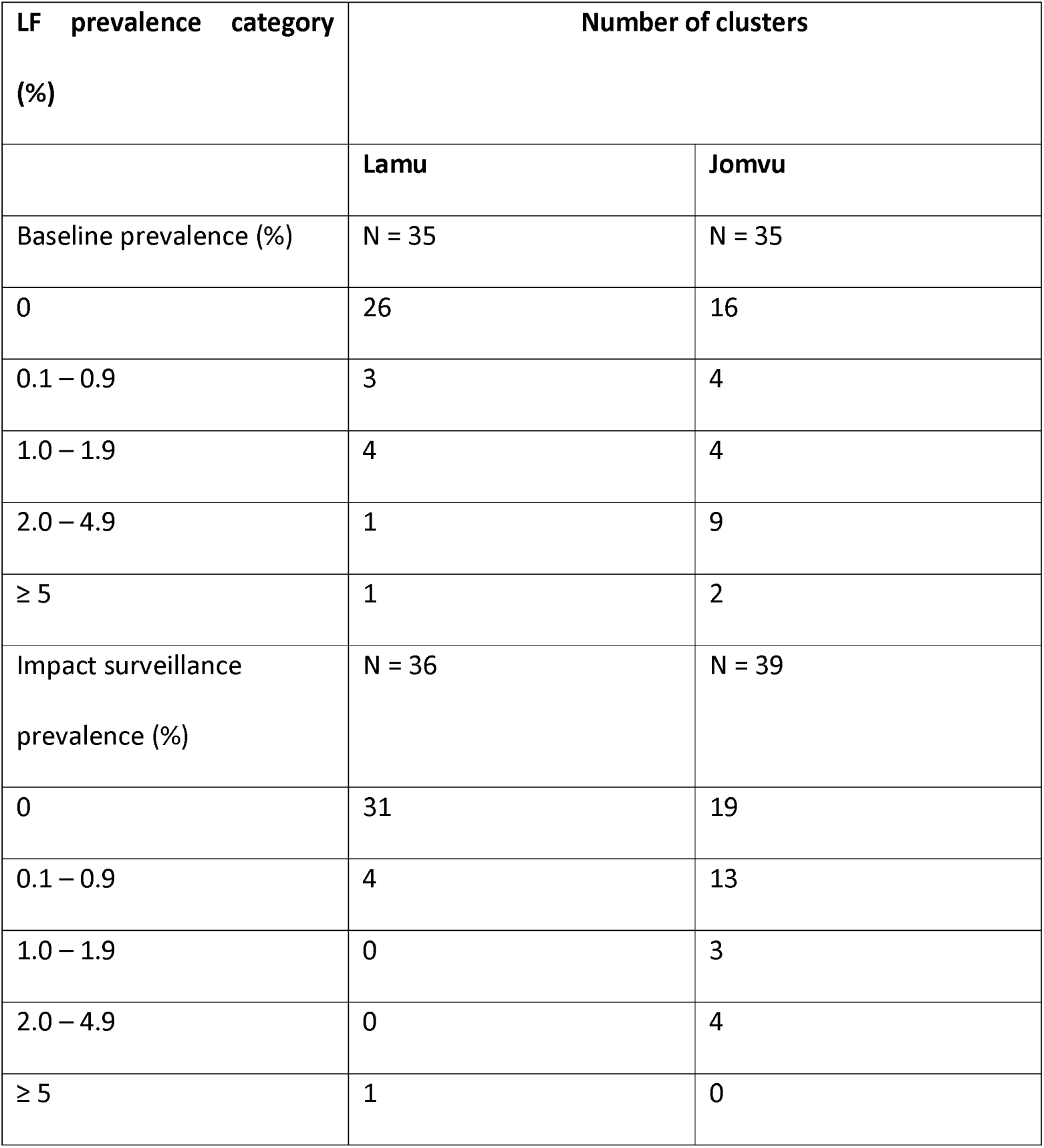
LF positivity among the study clusters (villages) in the two EUs during the baseline and endline surveys stratified by point prevalence categories.

At the endline survey, none of the 5–9-year-old children tested for LF infection were positive in Lamu, while 3 were found to be positive in Jomvu. Compared with the WHO recommended transmission survey (TAS), the FTS positive child cases observed in both EUs were below the threshold (Table 7).

**Table 7.** Endline IDA-MDA impact survey results in 5-9 years children, compared with transmission assessment survey (TAS) thresholds.

|  | Results during endline impact surveillance |  | WHO recommended TAS thresholds |  |
| --- | --- | --- | --- | --- |
|  | Jomvu | Lamu | Jomvu | Lamu |
| Number of clusters | 39 | 36 | 30 | 30 |
| Sample size | 1761 | 1647 | 1392 | 1532 |
| Cases/ critical cut-off | 3 | 0 | 16 | 18 |

The age group for the endline survey was children 5-9 years compared to the 6-7 years recommended for TAS, which increased the risk for failing TAS.

## Discussion

Compliance to the triple-drug regimen MDA in Lamu county and Jomvu sub-county was dramatically high due to excellent social mobilization by the National Programme for Elimination of LF (NPELF) and implementing partners. The various integrated strategies that were employed during MDA with IDA in the two areas in coastal region of Kenya showcased how a comprehensive approach can be employed to achieve high treatment coverage. Through partnerships, tailored communication, robust adverse event management, community engagement, innovative delivery mechanisms, effective CDD selection, and the involvement of LF champions, the program demonstrated the potential for achieving high treatment coverage. The treatment data demonstrated that the 80% treatment coverage target recommended by WHO could be realized. Indeed, the NPELF and partners were so successful in their social mobilization efforts that people from neighbouring sub-counties not meriting IDA-MDA travelled to Jomvu to receive the drugs (as is evident from the MDA coverage exceeding 100% in Jomvu). The results imply that NTD endemic communities will support the introduction of innovative treatments when the benefits of this change are clearly explained to them.

The results of the endline IDA-MDA impact survey offer strong evidence that it is safe to stop the IDA regimen in the two EUs because transmission appears to have been interrupted. Computer simulation modelling (14) predicts that in settings with a low baseline prevalence of LF (5%), the triple-drug regimen reduces the number of rounds of MDA required to reach the target prevalence by one or two rounds, compared with the two-drug regimen.

The detection of one Mf positive individual among the FTS positive cases in Jomvu indicates the presence of active LF transmission, albeit at relatively very low level and likely highly focal. In *W. bancrofti* endemic settings, demonstrating that the population prevalence of FTS positive cases is below a defined threshold in a transmission assessment survey (TAS) is an indication that LF is no longer a public health problem in the evaluation unit (15).

However, measuring CFA alone may not be an adequate way to for monitoring and evaluation of LF elimination programs using IDA-MDA (18). Indeed, evidence from initial IDA safety and efficacy clinical studies showed persistence of CFA for five years after a single round of IDA-MDA even with sustained clearance of Mf (10, 19). Thus, the development of new and more sensitive diagnostics tools, particularly for use during post-MDA with IDA surveillance, is urgently required to detect recrudescence. The use of entomological sampling and molecular detection of filarial DNA in mosquito vectors, known as molecular xenomonitoring is also considered a surveillance tool for LF that may have particular utility in low prevalence settings, and thus could supplement the standard diagnostics (20–22). However, although molecular xenomonitoring can also be used to detect the presence of the parasite in mosquito vectors and is a sensitive indicator of microfilaraemia, it is not a direct measure of infectivity or current rates of parasite transmission (15). Another challenge with molecular xenomonitoring is that it is not yet known how to interpret a signal of infection in mosquitoes (e.g., what is the threshold for action?) and the approach is laborious, expensive and requires diagnostic techniques that aren’t readily available to many NTD programs.

There is growing appreciation that basing MDA stopping decisions on the mean EU prevalence is not enough. As LF transmission approaches interruption, the remaining infection becomes increasingly focal and it is important to critically examine cluster-level results when determining whether a stop treatment decision is appropriate. The cluster data presented in Figs 1 and 2, as well as Table 4, present reassurance that post-IDA-MDA, there do not appear to be any remaining foci of infection that merit programmatic action. Nonetheless, the fact that 4 clusters in Jomvu and 1 in Lamu had a cluster-level FTS prevalence above 2% is an indication that the NPELF should remain vigilant of these sites during post-treatment surveillance to ensure that the lingering signal is a sign of waning infection and not recrudescence.

Since the implementation of MDA with IDA requires a substantial increase in programmatic resource expenditures, it may be necessary to restrict IDA-MDA to areas where the extra effort is necessary to drive transmission below the breakpoint. Consequently, it will be necessary to critically review the results of the transmission assessment surveys after the two rounds of MDA with IDA to determine if the EU meets the stopping criteria and can begin post-treatment surveillance. Intensive research surveys using serological approaches in American Samoa revealed residual hotspots with ongoing LF transmission several years after MDA was stopped (23, 24). Therefore, conducting an active post-treatment surveillance may be appropriate to determine if and when there is recrudescence.

Monitoring and evaluation (M&E) and surveillance are critical activities of any NTD control or elimination programme. The interim guidance for IDA-MDA impact surveillance is to test adults aged 20 years and above for circulating filarial antigenemia in 30 randomly selected clusters. The finding that there was significant reduction of LF infection prevalence among the adult population after the two rounds of annual IDA-MDA in the random clusters corroborates the proposal to conduct post-treatment surveillance among adults in randomly selected villages, and to employ microfilaraemia as an indicator for the presence of current infection. In an effort to make post-treatment surveillance of NTDs cost-effective in low-resource settings, a model-based geostatistics approach that would result in a survey design that requires sampling fewer clusters and a smaller total sample size while still achieving the same predictive qualities as conventional MDA impact surveillance approaches has been proposed (25, 26). It will therefore be important to consider conducting a follow-on operational research study as part of the second IDA-MDA impact surveillance in the two settings in order to provide additional evidence on the application of model-based geostatistics for its application in LF endemic countries where MDA with IDA is being considered.

This study has several limitations worth highlighting. The coast region of Kenya may be considered a low prevalence setting due to the impact of several rounds of the two-drug regimen MDA with DEC and albendazole and the widespread bednet usage. Lamu had received one round of MDA in 2011 (66% epidemiological treatment coverage) before the NPELF stopped mass treatments. Thereafter, Lamu received 3 rounds of consecutive MDA rounds in 2015, 2016 and 2017 and all were above the 65% epidemiological treatment threshold. Lamu was included primarily due to the historical persistence of LF infection in Lamu East particularly in Ndau island. With regard to Jomvu, two rounds of MDA with DEC and albendazole had been given in 2016 and 2017 but both were below the recommended 65% threshold. The missed opportunity to assess the impact of IDA-MDA on other diseases, particularly transmitted helminthiasis (STH), scabies, and head lice may be considered a study limitation. Ivermectin is known to act strongly against a wide variety of insects, nematodes, and acarine parasites, including lice and scabies (27, 28). A study to assess the impact of IDA-MDA for LF and STH control in Timor-Leste reported significant reduction in the prevalence of scabies, impetigo, and STH (*Trichuris trichiura* and *Ascaris lumbricoides*) among schoolchildren (29).

A noteworthy strength of the study, however, is that the design of the cross-sectional surveys provided for the inclusion of both children and adult populations in both the baseline and endline impact assessment. The random sampling in Jomvu at baseline revealed pockets of ongoing transmission hitherto unknown to the NPELF. The results of the study also indicate that purposive sites have a critical role in LF surveillance, particularly in identifying areas with highest risk of LF transmission (putative hot spots). In Lamu for example, Ndau island is historically known to be an area with relatively high LF transmission (30), which was also confirmed during a previous survey (8). Laboratory serologic and molecular (PCR) results are still forthcoming and when reported will shine even greater light on the impact of MDA with IDA – both spatially and across the age groups.

In conclusion, this study shows that the two rounds of IDA-MDA reduced LF infection below the recommended thresholds in the two EUs. The study also demonstrated that programmatic delivery of MDA with IDA is a feasible and efficient strategy for endemic countries looking to accelerate their LF elimination programs as they move towards post-treatment surveillance. The operational research study enabled baseline and endline collection of data that will be useful to guide global policy on monitoring and evaluation of LF programs that implement IDA-MDA. With a remaining, albeit very low, signal of CFA in the community, it is important to implement a post-treatment surveillance system which would enable efficient detection of any recrudescence of LF infection transmission at a sub-evaluation unit level.

## Supporting information captions

**S1 Table.** Sample size determination during the baseline survey conducted in 2018.

**S2 Table.** Sample size determination during the impact surveillance conducted in 2021.

## Data Availability

Data cannot be publicly shared because it contains potentially identifying information of human subjects. However, the data are available from the Scientific and Ethics Review Unit of Kenya Medical Research Institute for researchers who meet the criteria for access to confidential data.

## Acknowledgements

We express our gratitude to the county departments of health in Lamu and Mombasa for their excellent cooperation and support of the MDA with IDA and the associated operational research study. We would also like to thank the fieldwork teams for assistance with data collection. The local residents and operational research participants are acknowledged for their voluntary participation and commitment in this study. The partners, including Evidence Action, Africa Institute for Health and Development, Division of Vector-Borne and Neglected Tropical Diseases, Kenya Medical Research Institute, and the NTD Support Centre provided excellent technical support. The study was generously funded by the United States Agency for International Development (USAID) and Bill & Melinda Gates Foundation (BMGF) through the Coalition for Operational Research on Neglected Tropical Diseases (COR-NTD).

---

**S1 Table.** Sample size determination during the baseline survey conducted in 2018.

| **Survey Timepoint** | **Survey Area** | **Estimated Population** | **Sample size for random cluster survey of children aged 5-9 years** | **Sample size for random cluster survey of adults\*** | **Sample size for purposively selected sites of children aged 5-9 years** | **Sample size for purposively selected sites of adults\*** |
| --- | --- | --- | --- | --- | --- | --- |
| Baseline | Lamu | 144,000 | 1380 | 1710 | 200 (40 per site x 5 sites) | 200 (40 per site x 5 sites) |
|  | Jomvu | 140,000 | 1380 | 1710 | 200 (40 per site x 5 sites) | 200 (40 per site x 5 sites) |
| Endline | Lamu | 144,000 | 1380<br>(average of 46 kids per site) | 3150 (an average of 105 adults per site) | 50 per site | 100 per site |
|  | Jomvu | 140,000 | 1380<br>(average of 46 kids per site) | 3150 (an average of 105 adults per site) | 50 per site | 100 per site |

\*During baseline adults were considered as participants aged 10 years, during endline adults
were considered as participants aged 18 years

**S2 Table.**
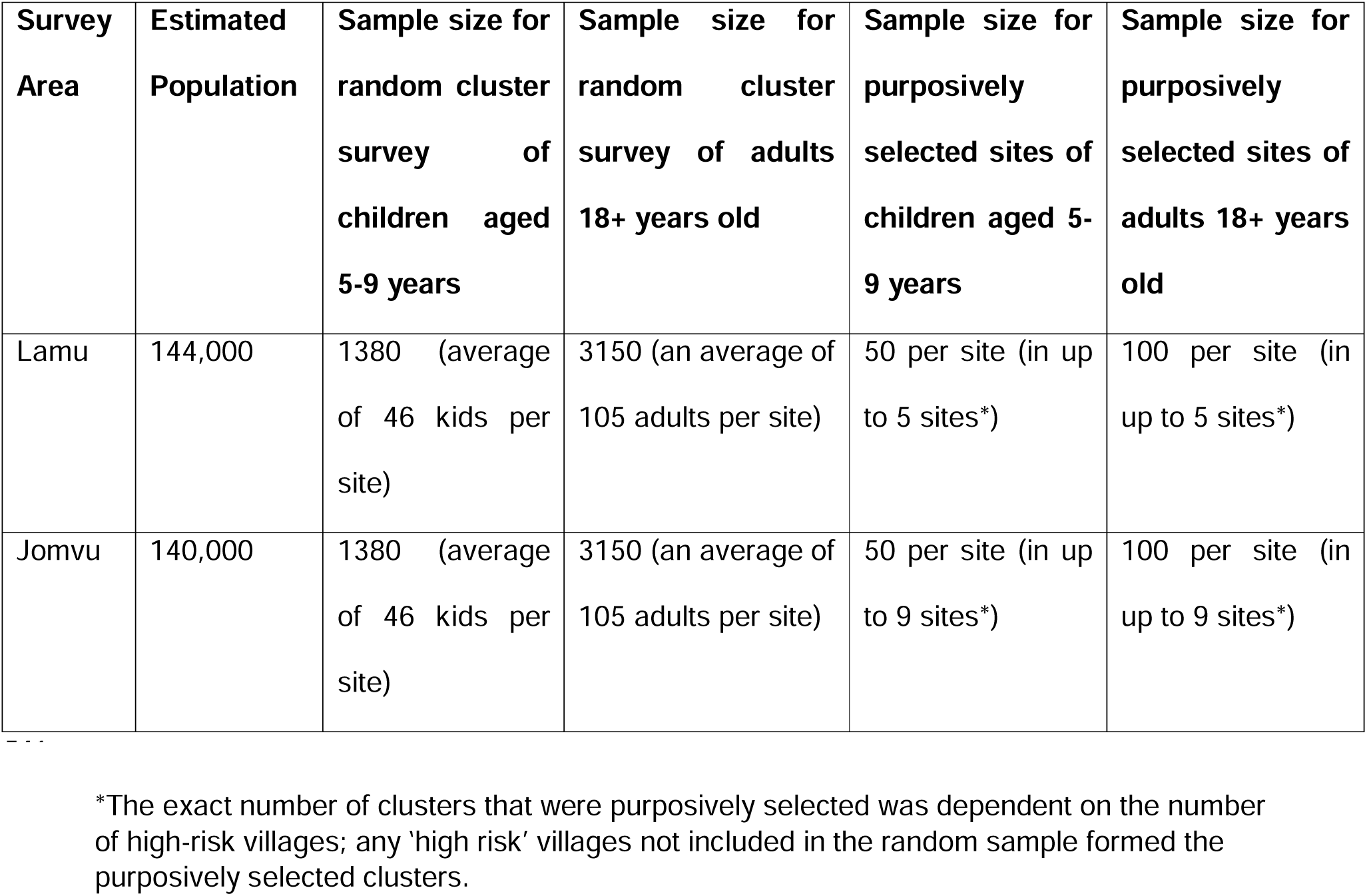
Sample size determination during the impact surveillance conducted in 2021.

## Notes

### Competing Interest Statement

The authors have declared no competing interest.

### Funding Statement

This work received financial support from the Bill & Melinda Gates Foundation
through their support of the Coalition for Operational Research on Neglected Tropical
Diseases (COR-NTD) grant. COR-NTD is funded at The Task Force for Global Health
primarily by the Gates Foundation and the United States Agency for International
Development (USAID). The funders had no role in study design, data collection and analysis, decision to publish, or preparation of the manuscript.

### Author Declarations

Ethical approval of the study protocols and informed consent documents was obtained from the Kenya Medical Research Institute Scientific and Ethics Review Unit (KEMRI/SERU Protocol No. 3721 and 4099).

